# The Effectiveness of aromatherapy and its supportive Interventions on anxiety and pain among breast cancer patients: A systematic review and meta-analysis

**DOI:** 10.64898/2026.06.16.26355762

**Authors:** Nathanaens O. Estervil, Cheryl L. Carmichael, Garumma T Feyissa

## Abstract

**Introduction:** Breast cancer treatments are often associated with pain and anxiety, which can hinder physical functioning and overall quality of life, even after treatment. Complementary therapies, such as aromatherapy, can be used to alleviate pain and reduce anxiety in breast cancer patients. This project aimed to synthesize current global evidence on the effectiveness of aromatherapy.

**Method:** This systematic review followed the PRISMA 2020 guidelines, with a comprehensive, systematic search conducted in PubMed, CINAHL, Cochrane Library, and SCOPUS for randomized controlled trials (RCTS) published from 2015 to 2025. Eligible studies included adult women breast cancer surgery patients who received aromatherapy during various periods of breast cancer. Where possible, data from the included studies were pooled using meta-analysis. GRADE approach was used to assess certainty of findings.

**Results:** The search yielded 84 studies. Out of these, six were included in this review. On average, aromatherapy reduces pain and anxiety scores by 0.79 (standard mean difference (SMD)=-0.79, 95% CI -1.42, -0.16) and 0.53 (SMD=-0.53, 95 CI=-0.90, -0.16) units, respectively, compared to control condition [Low-quality of evidence]. The combination of aromatherapy with music reduces pain and anxiety by 1.26 (SMD= -1.26, 95 CI=-1.65, -0.87) and 1.08 (SMD = -1.08, 95 % CI: -1.45, -0.70) units respectively compared to standard care [Low-quality of evidence].

**Conclusion:** There is a potential role for the use of aromatherapy and music therapy, to alleviate anxiety and pain, especially for non-preoperative anxiety and pain. Further research is needed to inform the integration of aromatherapy into the management of anxiety and pain.

## Introduction

Breast cancer is the most prominent, diagnosed cancer among women globally, remaining the second most prevalent cancer in the world(1). It once was the leading cause of cancer death in 103 countries across the world with incidence rates far exceeding those for other cancers (2). Due to advancements in early detection, technology and medical treatments, survival rates have significantly improved. Various breast cancer treatments are used depending on the type of breast cancer, the stage, and the person’s overall health. These treatment options often include a combination of treatment methods such as surgery (lumpectomy, mastectomy), systemic therapies (chemotherapy, immunotherapy, hormone therapy, etc.) and reconstruction (3).

Despite the improvement in the surgical techniques and treatments offered for breast cancer, treatment-related side effects are increasingly becoming an issue, lowering the quality of life for these patients. Women undergoing breast cancer surgery and/or treatment often experience pain, nausea, vomiting, fatigue, stress, anxiety and other various side effects (4). Amongst these side effects, perioperative anxiety has been shown to amplify the intensity of pain (5). Anxiety occurs due to the fear of death and/or poor quality of life which results in a feeling of a loss of control among cancer patients (6). At heightened, severe levels, the psychological and physiological effects of anxiety can lead to sensations of pain-the sensory and emotional experience of actual and/or potential tissue damage (7). This relationship between anxiety and pain management is essential to consider in the recovery of women undergoing breast cancer treatments, especially surgery (8). This causal relationship between these symptoms and breast cancer treatments calls for an intervention that promotes the effective management of physiological and psychological symptoms.

Non-pharmacological approaches are increasingly being considered as a complementary option, alongside standard treatment, for the symptom management of breast cancer treatment-related side effects (9). These approaches can be referred to as complementary and alternative medicine therapies (CAM)- treatments used alongside standard care (10). Emerging evidence from systematic reviews highlights the effectiveness of CAM treatments such as music therapy, massage, acupuncture, guided imagery, and aromatherapy in reducing anxiety and pain (8). For instance, a systematic review of randomized controlled trials (RCTs) and quasi-experimental studies in women undergoing breast cancer reported that music and acupuncture were associated with moderate reductions in pain and aromatherapy had a similar effect on perioperative anxiety (8). This exemplifies the growing interest in non-pharmacological and complementary interventions that may potentially be integrated into standard care to holistically manage breast cancer and its treatment related side effects.

Aromatherapy is one of the complementary interventions that has gained attention across studies within the field of medicine, specifically oncology. This intervention is a holistic practice, well known for the medical use of plant extracts, called *essential oils*, with antibacterial and healing properties (11). Essential oils such as lavender, lemon and bergamot are unique in their scents, their use, and effects. These essential oils, sometimes referred to as volatile oils, are extracted from various parts of the plant: petals, barks, stems, leaves, roots, fruits, etc. The methodology of aromatherapy stems from their antiseptic and skin permeability properties which allows for inhalation, local application and baths (12).

Growing evidence indicates that aromatherapy can effectively alleviate symptoms, such as pain and anxiety, in addition to nausea, vomiting and sleep disturbances for cancer patients (11). Lavender, specifically, can act as a sedative for wound healing and reducing sensations of pain and anxiety (11). The physiological mechanism behind this sensory experience centers around the olfactory nerve, connected to the limbic system-the emotional center of the human brain that plays an influential role in stress responses and pain perception (13). Furthermore, the ability of essential oils to penetrate the subcutaneous tissue or to be integrated into the biological signaling of receptor cells in the nose are essential to the delivery of this therapy (12). Based on these advantages and benefits of these neural stimulations, aromatherapy has been suggested as a potential effective intervention that can impact the psychological and physiological experience of breast cancer patients.

Few systematic reviews have examined the effectiveness of non-pharmacological interventions for symptom management in breast cancer patients, often including aromatherapy (8, 14, 15). In fact, aromatherapy is often combined with other complementary treatments such as music therapy and massage. However, there is limited synthesized evidence evaluating the effectiveness of aromatherapy alone. This has limited the ability to draw specific conclusions on the effectiveness of aromatherapy in managing treatment-related symptoms of breast cancer, specifically anxiety and pain.

Aromatherapy is one of the low-cost interventions to manage anxiety and pain. Despite the promising findings from preliminary search, evidence has shown mixed results. One study, for instance, has reported that aromatherapy had “no effect on cancer-related pain among breast cancer patients” (16). On the other hand, recent studies such as study by Zhang and colleagues have reported a significant effect of aromatherapy in reducing pain (17). These inconsistencies suggest that the therapeutic impact of aromatherapy may be influenced by contextual factors, such as timing of intervention, its delivery, or its integration with other supportive treatments. A systematic review is needed to understand the sources of such inconsistencies and the best available up-to-date evidence on the overall effect of aromatherapy across studies.

Given the growing emphasis on evidence-based, patient-centered and non-pharmacological approaches to breast cancer care, a critical synthesis of existing literature focused on aromatherapy and its combination with supportive modalities is warranted. There is no updated systematic review capturing all the recent research evidence on the effects of aromatherapy alone or its combination with complementary therapies in the context of reducing pain and anxiety levels of breast cancer patients throughout various stages of their cancer or phases of treatment.

Thus, this systematic review sought to clarify the extent at which aromatherapy is effective in reducing pain and anxiety. By providing a structured and comprehensive overview of the current evidence within the last ten years, this review aimed to contribute to the evidence base on aromatherapy and the potential of the integration of CAM therapies into the clinical care of breast cancer patients.

## Research questions

This review aimed to synthesize evidence on the effectiveness of aromatherapy, alone or in combination, in reducing pain and anxiety among breast cancer patients. Specifically, this review sought to determine:

a. The effectiveness of aromatherapy alone, to reduce anxiety among breast cancer patients;
b. The effectiveness of aromatherapy, in combination with other interventions, to reduce anxiety among breast cancer patients;
c. The effectiveness of aromatherapy alone, to reduce pain among breast cancer patients;
d. The effectiveness of aromatherapy, in combination with other interventions, to reduce pain among breast cancer patients.

## Methods

This systematic review was conducted and reported based on the Preferred Reporting Items for Systematic Reviews and Meta-Analyses (PRISMA) 2020 guidelines (18) (Supplementary file 1) and a priori protocol (CRD420261421918) accessed from https://www.crd.york.ac.uk/PROSPERO/view/CRD420261421918.

### Search strategy

A systematic search was conducted in the following databases: PubMed, CINAHL, The Cochrane Central Register of Controlled Trials (CENTRAL) and SCOPUS for randomized controlled trials (RCTS) published within the last ten years (2015–2025). The search strategy for each database is shown in Supplementary file 2. The following key words and their respective index terms were used: aromatherapy, breast cancer, breast neoplasms, pain, anxiety, RCTs (random controlled trials). The final search was conducted on the 12th of February 2026. To maximize our chances of identifying all relevant studies, in addition to searching databases, we also manually checked the reference lists of the included studies.

### Inclusion criteria

The inclusion criteria for the papers included in this review are as follows.

### Population

This review considered studies that focused on adult women diagnosed with breast cancer throughout various periods of their treatment.

### Intervention

This review considered studies that evaluated the effect of aromatherapy, operationalized as the use of essential oils through various routes of administration such as inhalation, topical application and steam bath. Studies in which aromatherapy was used as a standalone therapy or in combination with other interventions, such as music therapy, massage, yoga, etc. were included.

### Comparators

This review considered standard care, placebo effect or other alternative treatment(s) delivered by clinicians/health professionals or no interventions, as comparators.

### Outcome

The primary outcomes considered for this review were pain levels and anxiety symptoms measured qualitatively (presence/absence) or quantitatively (by different measures levels) using scales or validated assessment tools.

### Types of studies

All randomized controlled trials published in the English language were considered for inclusion. Pilot studies and feasibility studies were excluded from this review.

### Study selection

We collated the retrieved references using the Zotero citation software. After removing duplicates, the references were screened by title and the abstract for eligibility against inclusion by two reviewers. NOE and GTF independently screened the articles for inclusion. Discrepancies were resolved between the reviewers through discussion.

### Assessment of methodological quality

The methodological qualities of the eligible papers were assessed independently by two reviewers using the revised Joanna Briggis Institute (JBI) Critical Appraisal Tool, a standardized checklist for the assessment of risk of bias for randomized controlled trials.

### Data extraction and synthesis

Data were extracted using an a-priori developed data extraction tool. One reviewer extracted the data after which another reviewer checked and confirmed the accuracy of the data extracted. Whenever necessary, study authors were contacted. The data extraction tool contained study ID, country, sample size, intervention, outcome, and results at each of time points reported in the included papers. Data reported, in the form of mean and standard deviation, were extracted. Wherever summary values/effect sizes are missing, we calculated using the data provided for each intervention arm. After tabulating the intervention characteristics and the findings from each included paper, wherever appropriate, data on outcomes were pooled using meta-analysis using review manager (RevMan) software. We pooled and reported standard mean differences and their 95% confidence intervals. Since our primary interest was to draw conclusions beyond those studies included in this review, by default, we considered random effects model. However, random effects modeling is not generally recommended if the number of studies to be pooled is fewer than five (19–21). Therefore, we used fixed effects modeling whenever the studies included in the meta-analysis are fewer than five. We assessed the presence and severity of statistical heterogeneity across studies using the standard chi squared and I² statistic. We explored sources of heterogeneity using visual evaluation of the confidence intervals in forest plots, and subgroup analysis by subgroups by timing of measurement. Sensitivity analysis was conducted by changing the model (fixed effects versus random effects) to assess robustness of the analysis and no difference was observed across the models. Wherever assumptions for meta-analysis were not met, we presented the data narratively. While pooling the findings of primary studies, we used intention to treat analysis approach by including all the participants initially randomized participants as denominators of their respective initial assignment. We generated funnel plot to assess publication bias in the meta-analysis.

### Assessing certainty in the findings

We determined certainty of evidence and created a Summary of Findings (SoF) table using the Grading of Recommendations, Assessment, Development and Evaluation (GRADE) approach and GRADEpro GDT (22).

## Results

### Study selection and characteristics

The initial search identified 84 studies. After removing 16 duplicates, the remaining 68 studies were screened. 61 studies were excluded at title and abstract screening, leaving seven studies for full text screening. Among these, seven studies were assessed for eligibility using full text assessment. One study (23), did not meet the intervention criteria due to the lack of use of an essential oil. Hence, a total of six randomized controlled trials were included in this systematic review (Figure 1).

**Figure 1.**
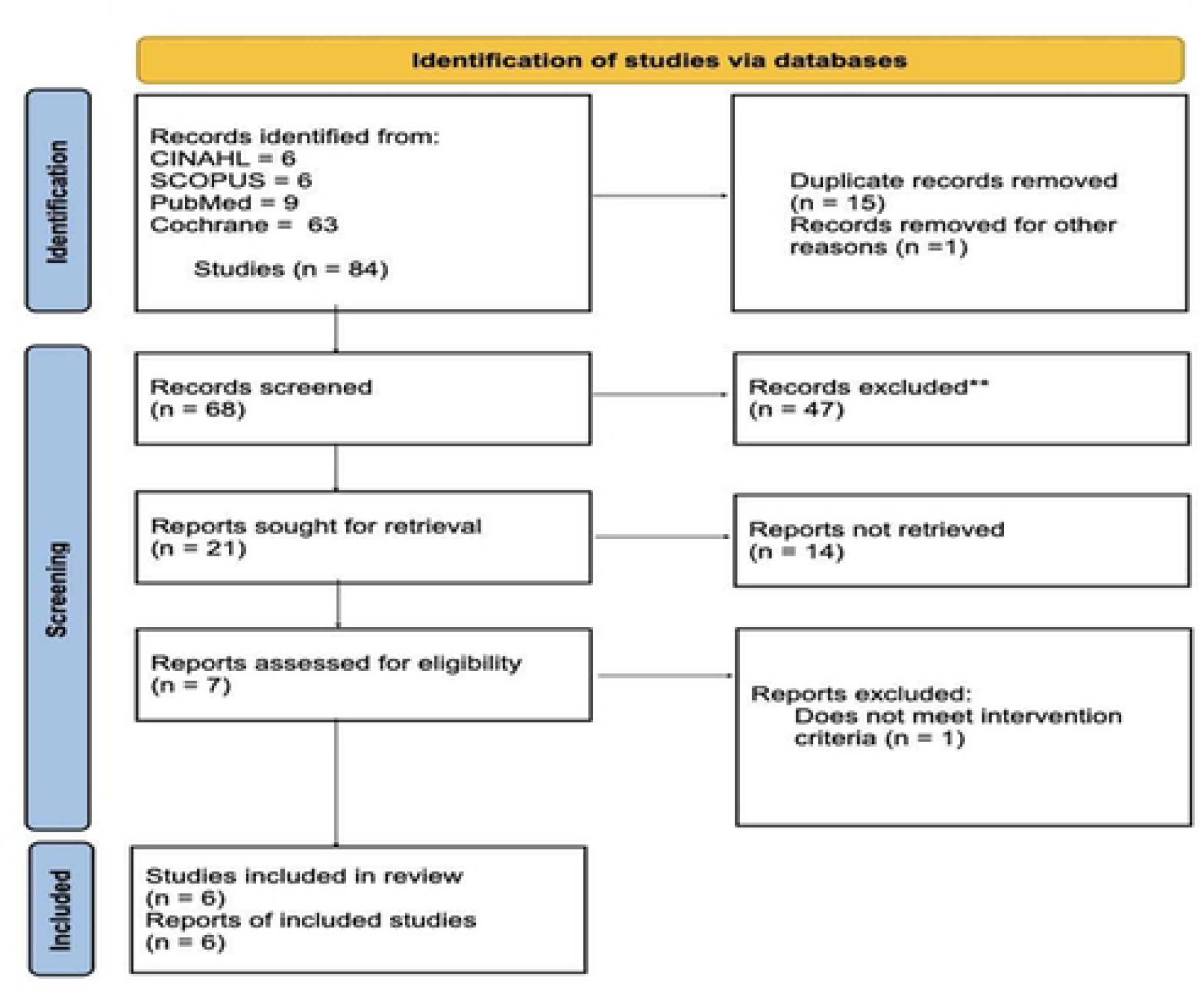
PRISMA Flow diagram (Source: Page MJ, et al. BMJ 2021;372:n71. doi: 10.1136/bmj.n71)

The included studies were conducted in Turkey, China, South Korea, Iran and the United States. The sample sizes of the studies ranged from 49 to 160 patients, with a total of 588 patients. The administration of aromatherapy varied across studies, including inhalation of essential oils throughout all studies that evaluated the effectiveness of aromatherapy alone and its combination with two supportive complementary alternative therapies-either music or massage. Lavender oil was the most frequently used essential oil, within four of the six (67%) studies (17, 24–26). One of these studies, (Deng et al., 2022 (25) included a mixture of lavender essential oil with two other essential oils-bergamot and geranium. The remaining two studies used frankincense essential oil (Ha et al., 2022 (27)) and rose essential oil (Samadi et al., 2025 (28)). Control conditions typically consisted of standard/usual care without aromatherapy or placebo, where coconut oil and sweet almond oil were used as the placebo oil, in the studies, Shammas et al., 2021 (26) and Zhang et al., 2025 (17), respectively **(Table 1).**

**Table 1.** Study Characteristics.

| Study ID | Country | Sample | Intervention/Control | Outcome |
| --- | --- | --- | --- | --- |
| Beyliklioğlu & Arslan, 2019 | Turkey | 80 patients scheduled for surgery:<br>Intervention (n=40)<br>control (n=40) | Intervention: Inhalation of 3-4 drops of lavender oil<br>Control: standard routine | Anxiety |
| Deng et al., 2022 | China | 160 patients scheduled for surgery:<br>MT (n=40)<br>AT (n=40)<br>CT (n=40)<br>UC (n=40) | AT: Mix of lavender, bergamot, and geranium oils<br>MT: Different music categories (classical, light, retro and popular)<br>CT: combination therapy<br>UC: usual care | Anxiety and Pain |
| Ha et al., 2022 | South Korea | 74 patients with acute pain syndrome who were to receive chemotherapy:<br>Group A (n=37)<br>Group B (n=37) | Intervention: ALT (massage) with frankincense essential oil and sweet almond carrier oil<br>Control: standard care for analgesia (acetaminophen 650/75 mg). | Pain |
| Samadi et al., 2025 | Iran | 99 female mastectomy candidates:<br>aromatherapy (n=33)<br>music therapy (n=33)<br>control group (n=33) | Intervention: inhalation of rose essential oil<br>Music therapy:<br>Control: standard care | Anxiety |
| Shammas et al., 2021 | USA | 58 post microvascular breast construction patients:<br>experimental group (n=29)<br>control group (n=29) | Intervention: Aromatherapy inhalation of lavender oil;<br>Control: Coconut oil (placebo) | Anxiety and Pain |
| Zhang et al., 2025 | China | 117 participants undergoing chemotherapy<br>Intervention group (n = 58)<br>Placebo group (n = 59) | Intervention: aromatherapy massage with lavender oil<br>Control: sweet almond oil over four weeks (placebo) | Anxiety and Pain |

**NB:** AT: aromatherapy, MT: music therapy, ALT- aromatic lymphatic tressage (massage)

**Table 2.** Appraisal table.

| Study ID | Beyliklioğlu & Arslan, 2019 | Deng et al., 2022 | Ha et al., 2022 | Samadi et al., 2025 | Shammas et al., 2021 | Zhang et al., 2025 |
| --- | --- | --- | --- | --- | --- | --- |
| Q1 | Y | Y | Y | Y | Y | Y |
| Q2 | U | U | U | U | U | Y |
| Q3 | Y | Y | Y | Y | Y | Y |
| Q4 | N | N | N | N | Y | Y |
| Q5 | N | N | N | N | N | Y |
| Q6 | Y | Y | Y | Y | Y | Y |
| Q7 | U | N | U | N | U | Y |
| Q8 | Y | Y | Y | Y | Y | Y |
| Q9 | Y | Y | Y | Y | Y | Y |
| Q10 | Y | Y | U | Y | Y | Y |
| Q11 | Y | Y | N | Y | N | N |
| Q12 | Y | Y | Y | Y | Y | Y |
| Q13 | Y | Y | Y | Y | Y | Y |
| Score | 9/13 | 9/13 | 7/13 | 9/13 | 9/13 | 12/13 |

Q1-Was true randomization used for assignment of participants to treatment groups? Q2- Was allocation to treatment groups concealed? Q3-Were treatment groups similar to the baseline? Q4-Were participants blind to treatment assignment? Q5-Were those delivering treatment blind to treatment assignment? Q6-Were treatment groups treated identically other than the intervention of interest? Q7-Were outcome assessors blind to treatment assignments Q8-Were outcomes measured in the same way for treatment groups? Q9-Were outcomes measured in a reliable way Q10-Was follow up complete and if not, were differences between groups in terms of their follow up adequately described and analyzed? Q11-Were participants analyzed in the groups to which they were randomized? Q12-Was appropriate statistical analysis used? Q13-Was the trial design appropriate, and any deviations from the standard RCT design (individual randomization, parallel groups) accounted for in the conduct and analysis of the trial.

### Methodological quality of included studies

Overall, the scores of the included randomized controlled trial studies ranged from seven to twelve out of thirteen JBI critical appraisal criteria. The main methodological limitations identified were low sample sizes, and lack of blinding. Some appraisal criteria were frequently reported as unclear, or “U”, due to insufficient methodological detail; in particular, information regarding allocation concealment, blinding of outcome assessors, and attrition management was not consistently described.

Four of the six studies (67%), Beyliklioğlu & Arslan, 2019; Deng et al., 2022; Samadi et al., 2025; Shammas et al., 2021,(24–26, 28), demonstrated high methodological quality by meeting nine out of the thirteen JBI criteria. The studies included in this study more commonly lacked blinding of participants due to the nature of the trials conducted. Samadi et al.,2025(28) also reported an open-label randomized controlled design, increasing the possibility of participant and researcher expectation bias. Furthermore, another study, Ha et al., 2022 (27) demonstrated the lowest methodological quality, scoring seven out of the thirteen JBI criteria. Ha at al., 2022 (27) conducted a randomized crossover trial evaluating aromatic lymphatic tressage- a combination of aromatherapy and lymphatic massage therapy. Furthermore, the study provided limited information on blinding (27). The lack of blinding and concealment in addition to the crossover design justifies its lower JBI criteria score. On the other hand, Zhang et al., 2025 (17) scored twelve out of thirteen JBI criteria, achieving the highest appraisal score among the included studies. This study employed a prospective double-blind, randomized placebo-controlled design, which minimizes both performance and detection bias **(Tabe 2).**

### Findings reported in the included studies

#### Pain outcomes

Four of the six studies, Deng et al., 2022 (25); Ha et al., 2022 (27); Shammas et al., 2021 (26); Zhang et al., 2025(17), reported on the effectiveness of aromatherapy on pain (Table 3). Among the four studies, two studies (50%) reported significant pain reductions following aromatherapy intervention. These two studies (Deng et al., 2022 (25) and Zhang et al., 2025(17)) reported on the effectiveness of aromatherapy combined with other CAM interventions, music and massage, respectively. The randomized controlled trial done by Deng et al., 2022 (25) found that patients receiving aromatherapy, either alone or combined with music, reported lower pain scores compared to those who only received usual care. Furthermore, the combination therapy (aromatherapy + music) showed greater improvement in pain scores compared to the administration of aromatherapy alone, which reported a change of 1.75 ± 1.06 (combination) vs. 3.03 ± 1.2 (alone). Zhang et al., 2025(17) similarly demonstrated significant reductions in pain scores among patients receiving aromatherapy massage with lavender oil compared to the placebo group with sweet almond oil (6.66 ± 1.45 vs 7.41 ± 1.19, respectively, p = 0.006).

**Table 3.** Pain outcomes.

| Study ID | Participants in each group | Results | Summary |
| --- | --- | --- | --- |
| Deng et al., 2022 | 160 patients:<br>MT (n=40);<br>AT (n=40);<br>CT (n=40)<br>UC (n=40) | T1 (30 min before surgery)<br>CT: $0.28 \pm 0.45$<br>AT: $0.35 \pm 0.62$<br>MT: $0.33 \pm 0.62$<br>UC: $0.50 \pm 0.72$<br>T2 (4 hrs after tracheal extubation)<br>UC: $6.13 \pm 1.02$<br>AT: $3.38 \pm 0.90$<br>MT: $3.53 \pm 0.72$<br>CT: $2.03 \pm 0.83$ | Patients receiving aromatherapy, either alone or combined with music, reported lower pain scores compared to those who only received usual care<br>Combination therapy (aromatherapy + music) showed greater improvement in pain scores compared to the aromatherapy alone ($p < 0.001$). |
| Ha et al., 2022. | 74 patients:<br>Group A (n=37);<br>Group B (n=37). | Peak Score<br>Group A (Standard care)<sup>a</sup><br>Cycle 1: 5.44 (2.64)<br>Cycle 2: 5.28 (2.45)<br>Group B (ALT)<sup>b</sup>:<br>Cycle 1: 5.82 (2.16)<br>Cycle 2: 5.05 (2.56)<br>Average Pain score<br>Group A (Standard care)<br>Cycle 1: 1.80 (1.73)<br>Cycle 2: 1.92 (1.76)<br>Group B (ALT)<br>Cycle 1: 1.94 (1.45)<br>Cycle 2: 1.73 (1.37).<br>Peak pain across cycles<br>ALT+ standard: 5.05 (2.56)<br>Standard: 5.28 (2.45)<br>Average score<br>ALT+ standard: 1.73 (1.37)<br>Standard: 1.92 (1.76) | Across both cycles, pain scores were lower in the intervention (ALT) group compared with the control (taxane) group after aromatherapy ($p = 0.36$ for peak score and $p = 0.216$ for average score). |
| Shammas et al., 2021 | 58 patients:<br>Experimental group (n=29);<br>Control group (n=29) | Pre-operative<br>Control: 0.5 (1.36)<br>Intervention: 0.8 (1.60)<br>POD 1<br>Control: 3.5 (1.91)<br>Intervention: 3.1 (2.74)<br>POD 2 (final)<br>Control: 2.5 (1.92)<br>Intervention: 2.9 (2.81)<br>($p = 0.3$) | No significant differences in pain scores between AT (lavender oil) and placebo (coconut oil) ($p = 0.30$). |
| Zhang et al.,<br>2025 | 117 participants:<br>Intervention group (n = 58);<br>Placebo group (n = 59). | (T1) First Week<br>IG: 4.66 (2.20)<br>CG: 6.43 (1.77)<br>(T2) Second week<br>IG: 5.66 (1.48)<br>CG: 6.88 (1.63)<br>(T3) Third week<br>IG: 6.45 (1.47)<br>CG: 6.75 (1.13)<br>(T4) Fourth Week<br>IG: 6.66 (1.45)<br>CG: 7.45 (1.19). | Significant reduction in pain score following aromatherapy compared to placebo group (p = 0.006). |

**NB:** VAS: Visual Analog Scale was used to measure pain level, AT: aromatherapy, MT: music therapy, ALT- aromatic lymphatic tressage (massage). <sup>a</sup>Group A received standard-of-care comprised of acetaminophen/tramadol in the 1st cycle and received additive ALT in the 2nd cycle; <sup>b</sup>Group B [ALT] received additive ALT in the 1st cycle and received standard-of-care only in the 2nd cycle.

However, the other two studies reported limited or non-significant effects of aromatherapy alone on pain. Ha et al., 2022 evaluated aromatic lymphatic tressage (ALT) using frankincense essential oil compared to standard treatment of taxane administration, which is standard for analgesia. Results showed slightly lower pain scores in the intervention (ALT) group compared with the control (taxane) group after aromatherapy administration (1.73 vs 1.92), although this difference was not statistically significant (p = 0.216). However, the study found that breast cancer patients who previously experienced TAPS (taxane acute pain syndrome) reported lower visual analogous scale (VAS) pain scores after receiving ALT. Similarly, Shammas et al., 2021 reported no significant differences in pain scores between patients receiving lavender oil and those receiving placebo, coconut oil (p = 0.30), which can be justified by the use of coconut oil in the comparison group. Nonetheless, the study reported the observation of significant differences in pain scores based on time, signifying improvement in pain scores throughout the length of the patients’ hospitalization Shammas et al., 2021 (26) (**Table 3).**

#### Anxiety outcomes

Five of the six included studies reported on the effectiveness of aromatherapy on anxiety (Beyliklioğlu & Arslan, 2019 (24); Deng et al., 2022 (25); Samadi et al., 2025 (28); Shammas et al., 2021 (26); Zhang et al., 2025 (17)). Among the five studies, four studies (80%) reported significant reduction in anxiety following aromatherapy interventions (Beyliklioğlu & Arslan, 2019 (24); Deng et al., 2022 (25); Samadi et al., 2025 (28); Zhang et al., 2025 (17)). Of these studies, four (80%) ((Beyliklioğlu & Arslan, 2019 (24); Deng et al., 2022 (25); Shammas et al., 2021 (26); Zhang et al., 2025 (17)) found significant anxiety reduction from the use of lavender oil in the administration of aromatherapy. Shammas et al., 2021 (26) found no significant differences in anxiety levels between the aromatherapy and placebo groups before and after surgery when measured using the Hospital Anxiety and Depression Scale, showing no significance (p value = 0.82).

Four out of the five (80%) studies examined the effectiveness of aromatherapy alone on anxiety (Beyliklioğlu & Arslan, 2019 (24); Deng et al., 2022 (25); Shammas et al., 2021 (26) and Samadi et al., 2025 (28)). Among the four studies, three (75%) of them found significant reductions in anxiety from the use of aromatherapy alone. The study by Beyliklioğlu & Arslan, 2019 (24); found that lavender oil inhalation significantly reduced anxiety scores among breast cancer patients, with mean STAI (State-Trait Anxiety Inventory) scores decreasing from 43.00 ± 11.48 to 37.28 ± 9.93 (p = 0.003) while no significant change was observed in the control group (p >.109). Similarly, Samadi et al. (2022) reported significant reductions in anxiety among 99 female mastectomy candidates receiving aromatherapy with rose essential oil, with mean NVAAS (Numeric Visual Analog Anxiety Scale) scores decreasing from 6.27 ± 1.56 to 4.18 ± 1.15, while a low mean change was reported from the control group (-0.45 ± 0.83).

The other study that examined the effectiveness of aromatherapy alone on anxiety was the study done by Deng et al., 2022 (25) which was also one of the two studies out of the five total (40%) that examined the effectiveness of its combination with other supportive CAM interventions on anxiety among breast cancer patients. Aromatherapy alone resulted in a mean anxiety reduction score of 3.18 units (mean difference (MD) = −3.18 ± 1.48) on the Visual Analog Scale, while combination therapy of aromatherapy and music therapy produced the greatest improvement (MD= −3.98 ± 1.19), compared with minimal change in the usual care group (MD=−0.73 ± 0.75). However, music therapy yielded the greatest reduction in anxiety compared to the other two groups, slightly more than aromatherapy. Zhang et al., 2025 reported significantly lower anxiety scores in patients receiving aromatherapy massage with lavender oil compared with the perceived increasing scores from the placebo group (22.17 ± 7.15 to 20.26 ± 6.73) vs (21.02 ± 4.97 vs 24.10 ± 6.46), respectively **(Table 4).**

**Table 4.**
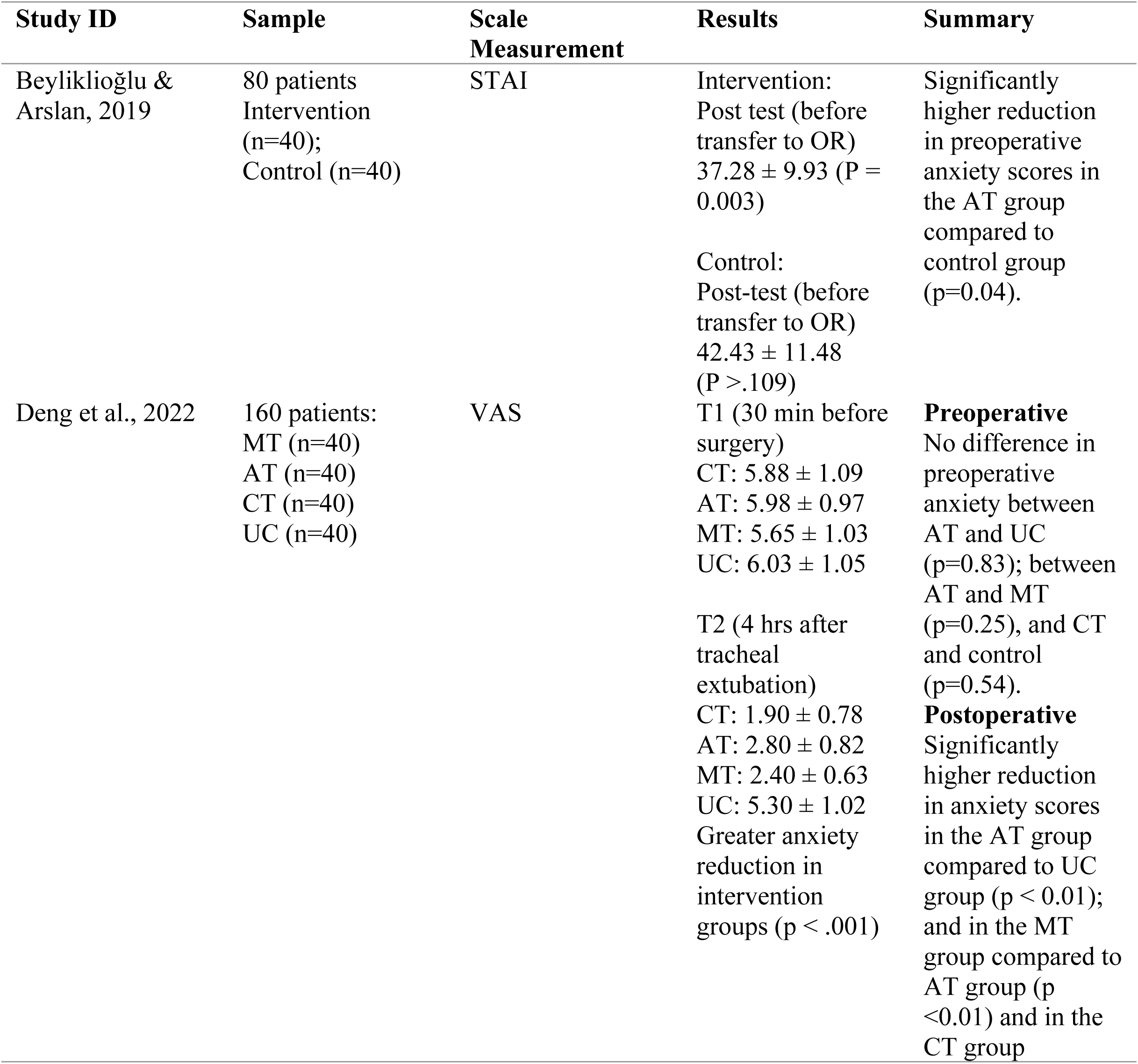
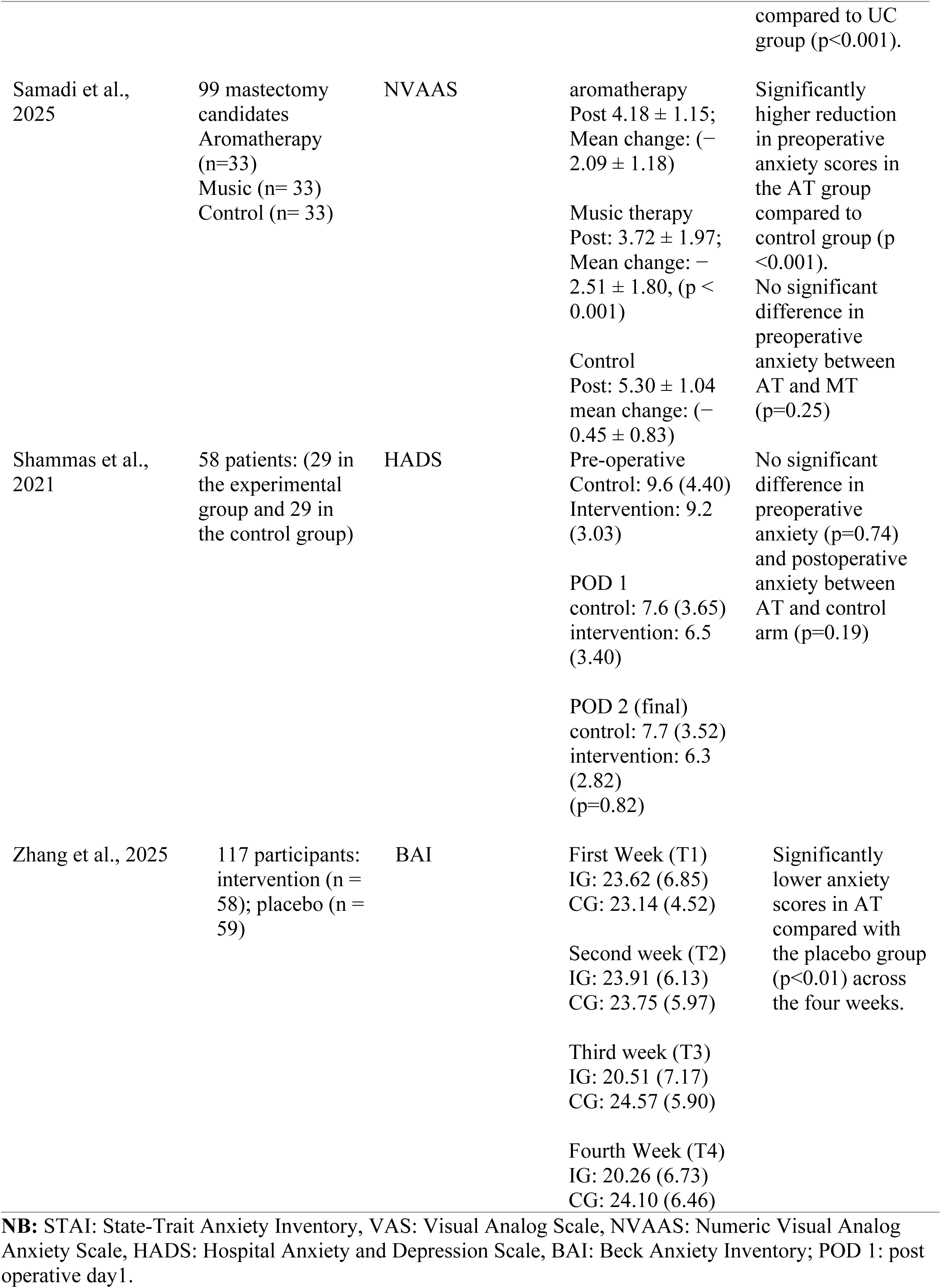
Anxiety outcomes.

### Pooled findings

#### Pooled effect of aromatherapy on pain

On average, aromatherapy reduced pain scores by 0.79 units (SMD=-0.79, 95% CI - 1.42, -0.16) compared to the control condition. However, this effect varies across time points. The effect of aromatherapy on preoperative pain is not statistically significant. The effect of aromatherapy on other time points varies across studies (Figure 2a). To assess the robustness of the analysis, we analyzed using fixed effect model, which produced slightly lower effect sizes (SMD=-0.63, 95% CI -0.77, -0.49) with no significant difference between the two models (**Figure 2b).** Most of the studies from which data for meta-analysis was pooled lacked blinding of participants, intervention administrators and outcome assessors due to the nature of the trials conducted. A visual inspection of funnel plot does not indicate any pattern of publication bias (**Figure 2c).** However, since the number of studies included in each analysis is less than 10, the analysis might not have been powered enough to detect significant publication bias (Afonso et al. 2024).

**Figure 2.**
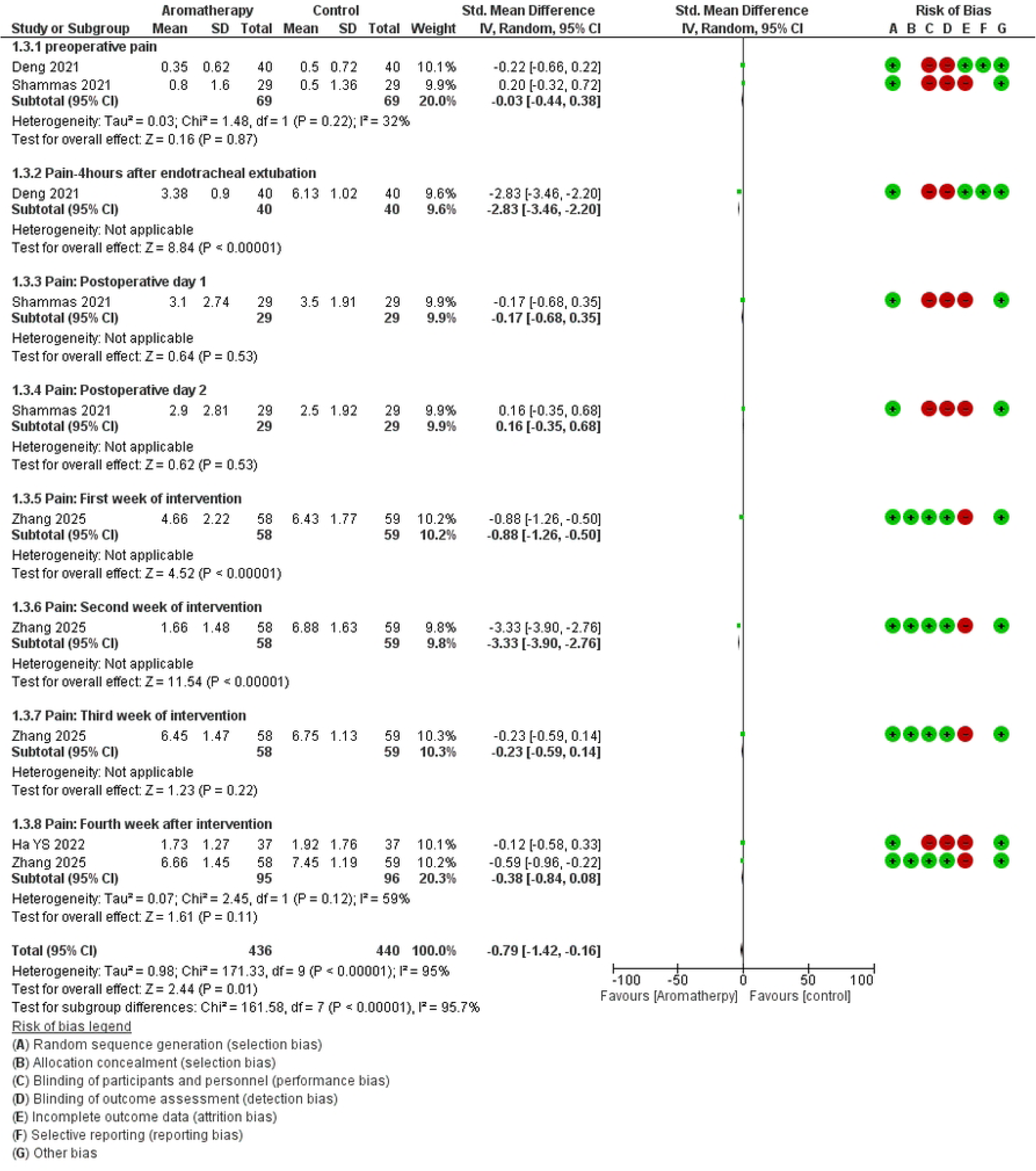
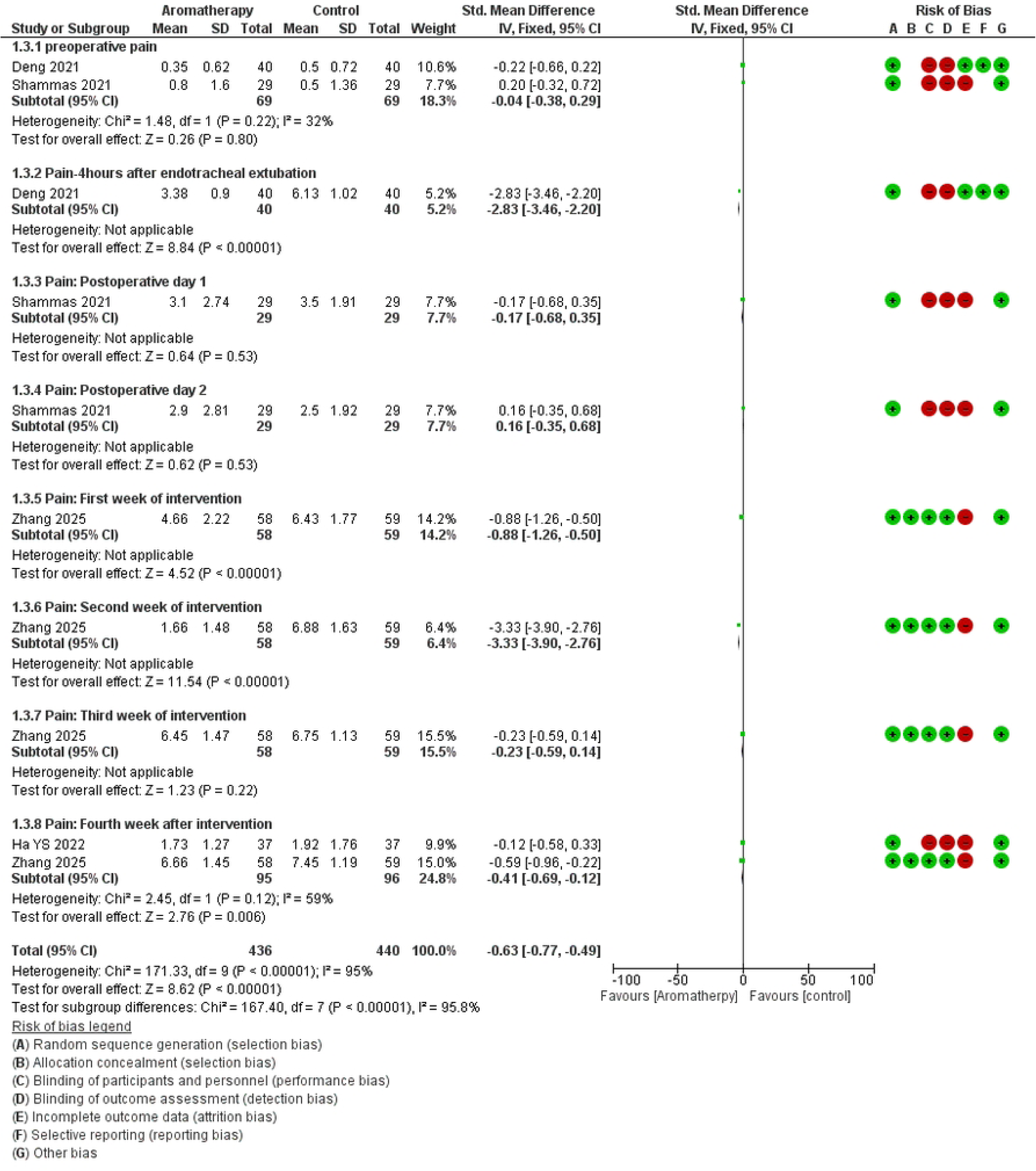
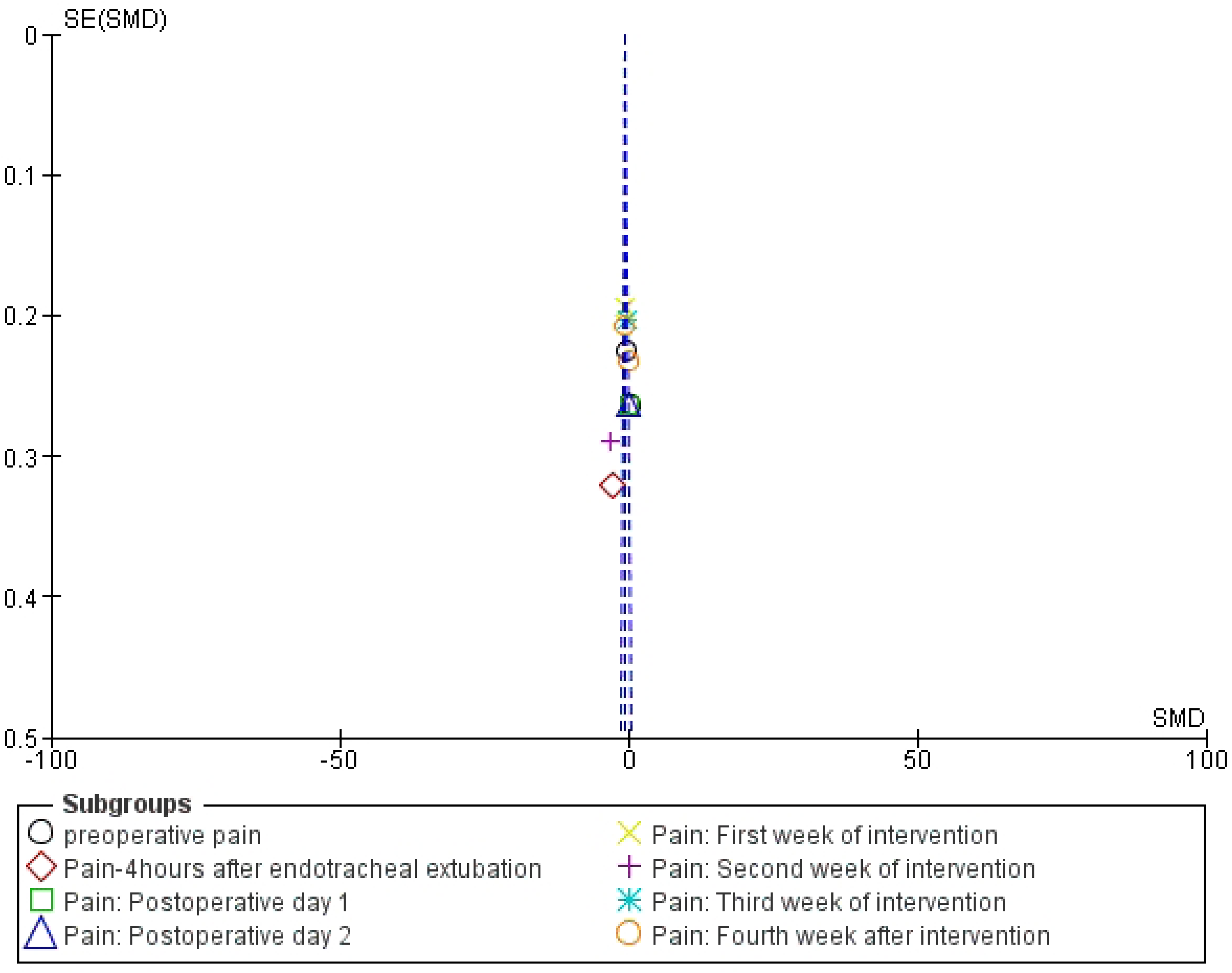
a. A forest plot showing the effect of aromatherapy on pain scores compared to usual care using fixed effects model.

The combined effect of aromatherapy and music therapy on preoperative pain is not statistically significant. Four hours after extubation of endotracheal tubes, patients exposed to the combined music and aromatherapy had a 4.37 (SMD=-4.37,95% CI -5.19, -3.55) units lower pain scores compared to the control condition. Overall, across the different time points, patients exposed to the combination of aromatherapy and music therapy had 1.26 (SMD= -1.26, 95 CI=-1.65, -0.87) units lower pain scores compared to usual care (**Figure 3).** There is no significant difference between the effect of aromatherapy and that of music therapy on both preoperative and postoperative pain scores (**Figure 4).**

**Figure 3.**
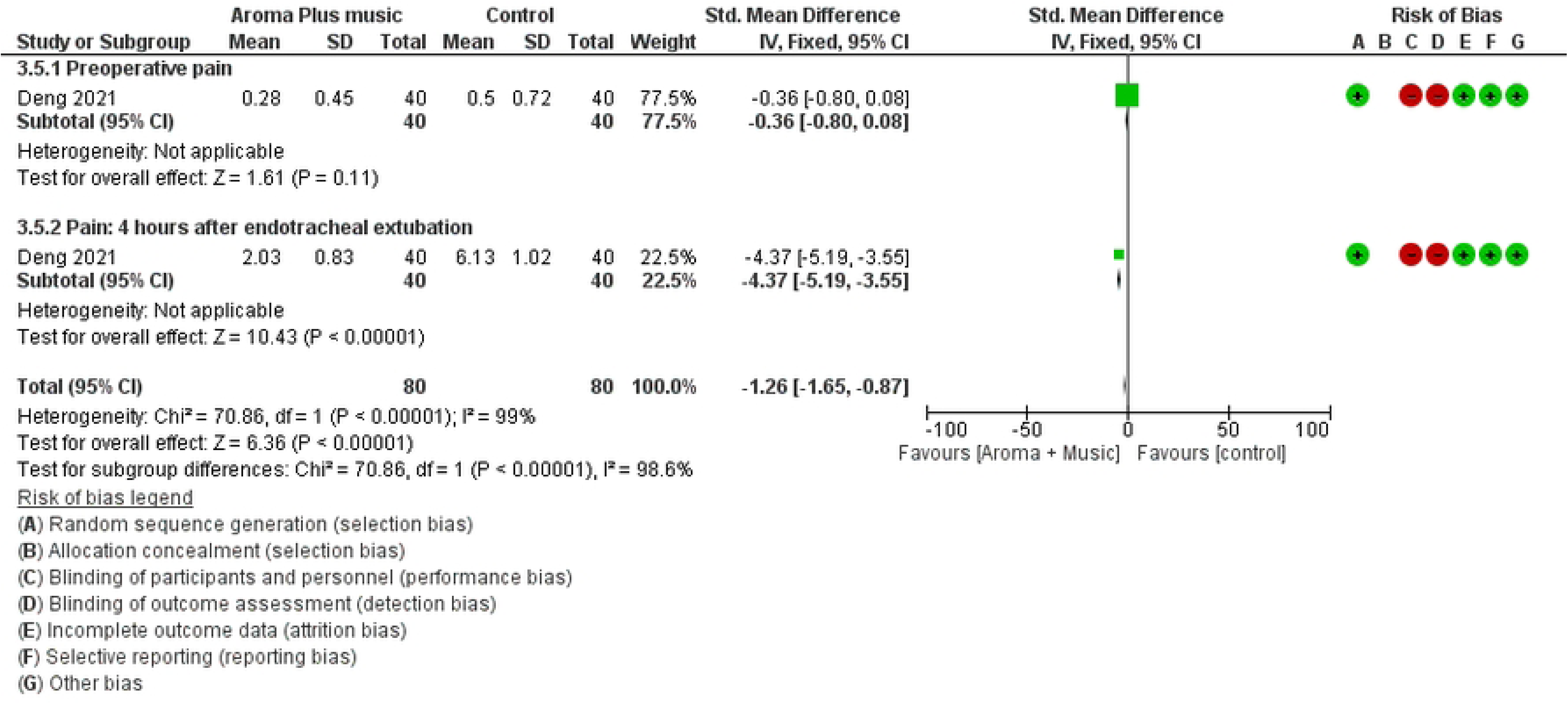
b. A forest plot showing the effect of aromatherapy on pain scores compared to usual care using random effects model.

**Figure 4.**
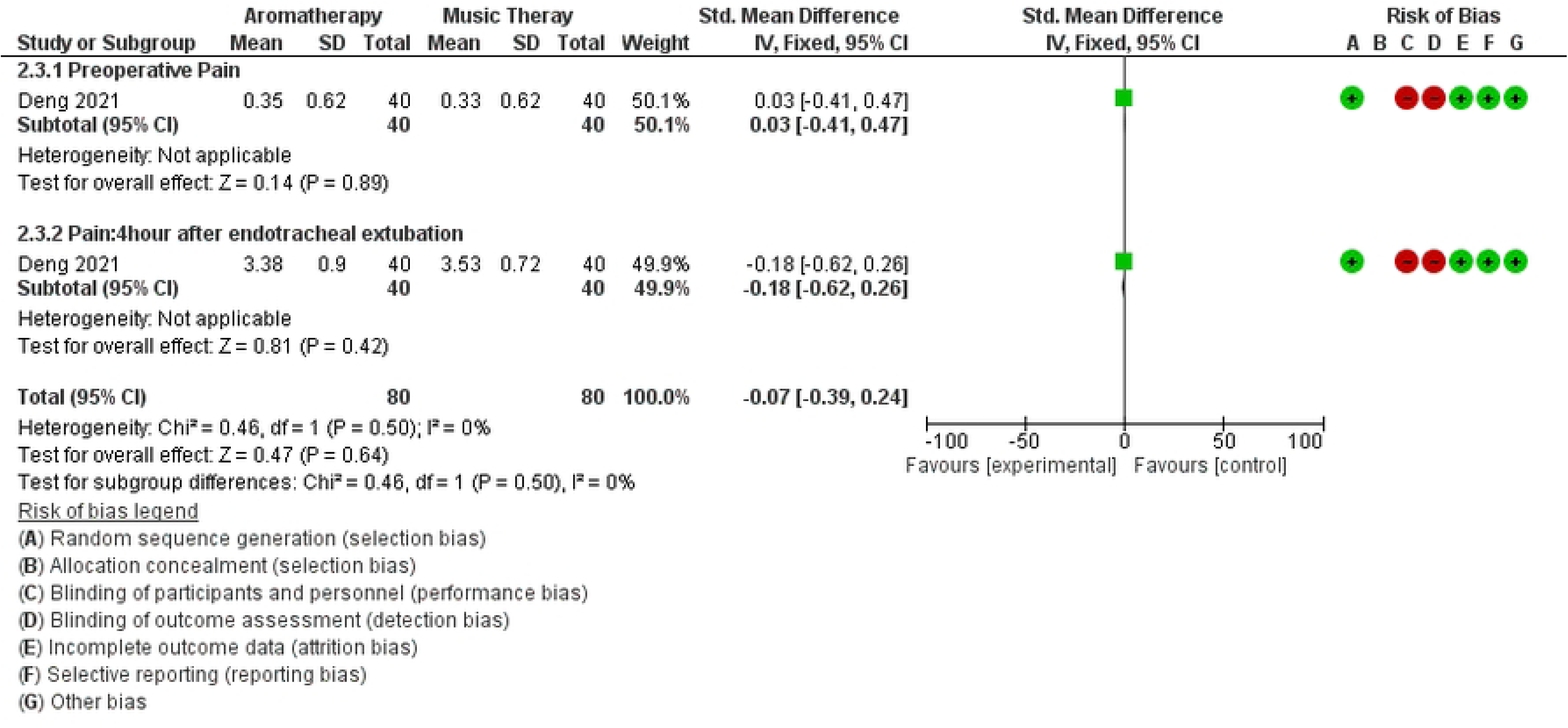
c. A funnel plot to assess publication bias for pain outcomes

#### Pooled effect of aromatherapy on anxiety

On average, aromatherapy reduced anxiety scores by 0.53 units (SMD=-0.53, 95 CI=-0.90, -0.16) compared to the control group. Considerable heterogeneity was observed (I^2^ = 87%; *p* < 0.01); hence we conducted subgroup analysis by timing of the measurements (**Figure 5a).** Fixed effect model showed slight decrement in effect size (SMD=-0.43, 95 % CI: -0.56, -0.30) with no change in the direction of effect (**Figure 5b).** The studies from which data for meta-analysis was pooled had average appraisal score of 9/13 and they commonly lacked blinding of participants, intervention administrators and outcome assessors due to the nature of the trials conducted. The funnel plot does not indicate any pattern of publication bias even though the limited number of studies included in the analysis is less than 10 and the power to detect bias is low (Afonso et al. 2024). **(Figure 5c).**

**Figure 5.**
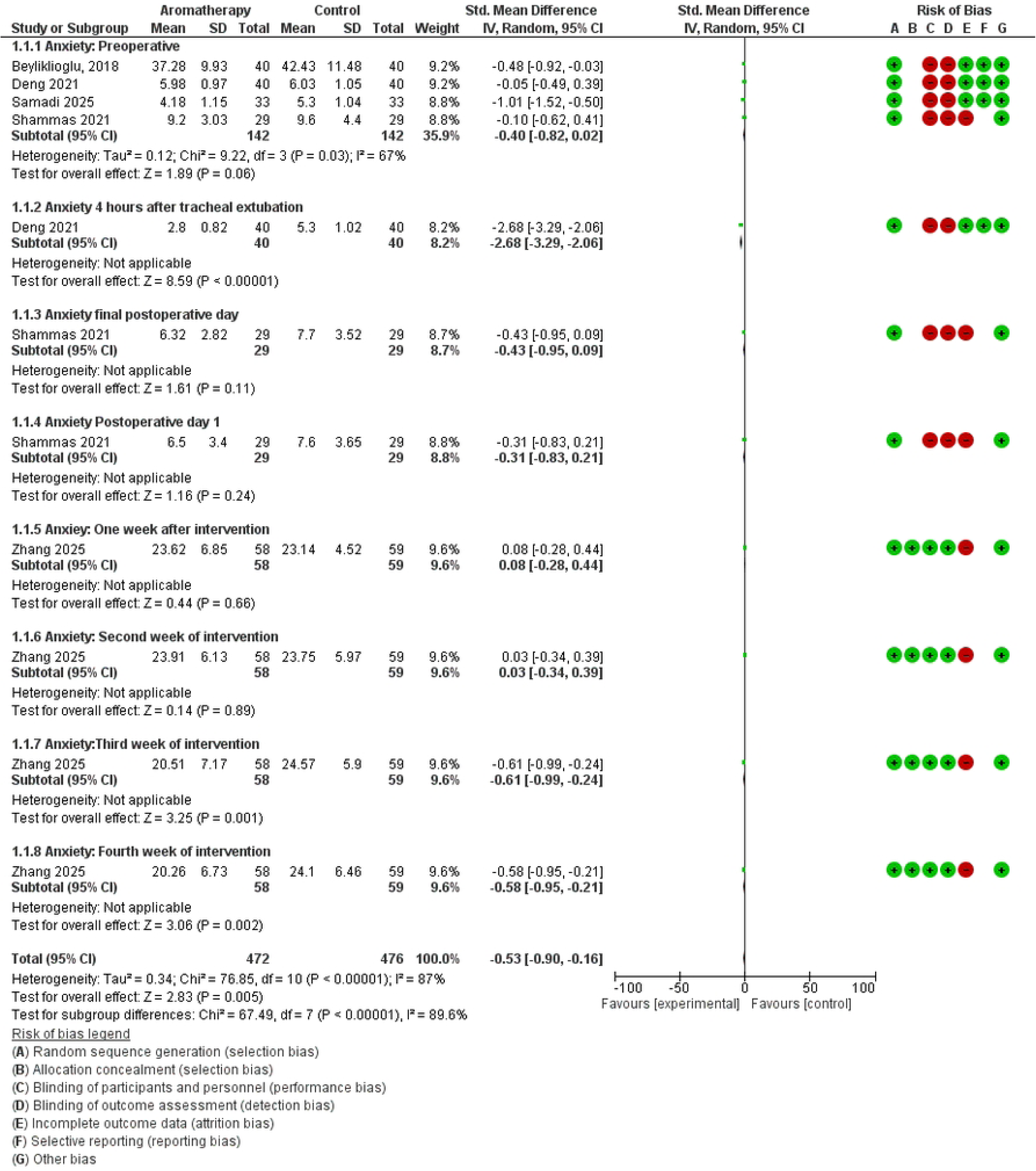
A forest plot showing the effect of aromatherapy combined with music on pain scores compared to usual care. **a.** A forest plot showing the effect of aromatherapy on anxiety scores compared to usual care using fixed effects model.

**Figure 5.**
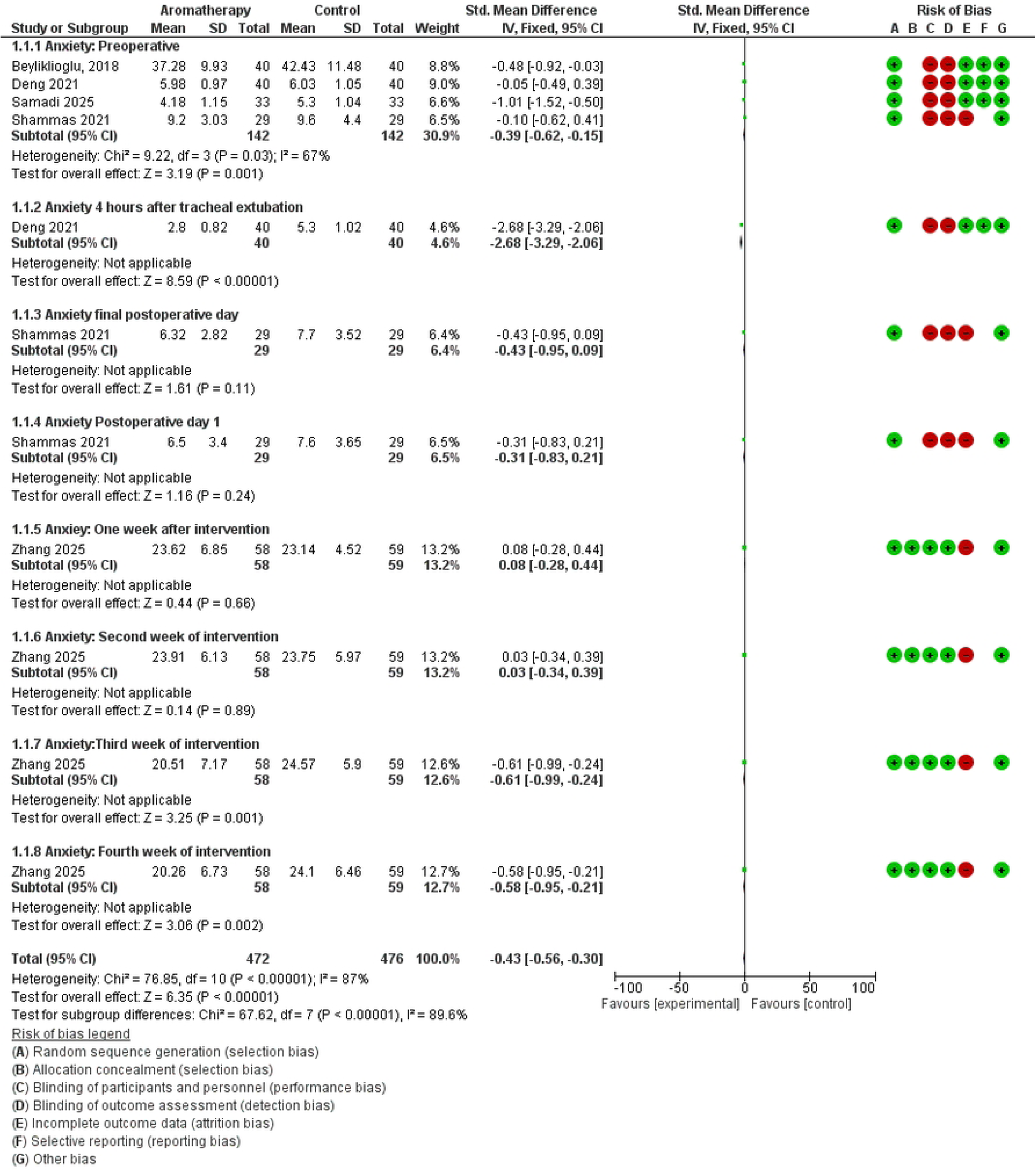
b. A forest plot showing the effect of aromatherapy on anxiety scores compared to usual care using random effects model.

**Figure 5.**
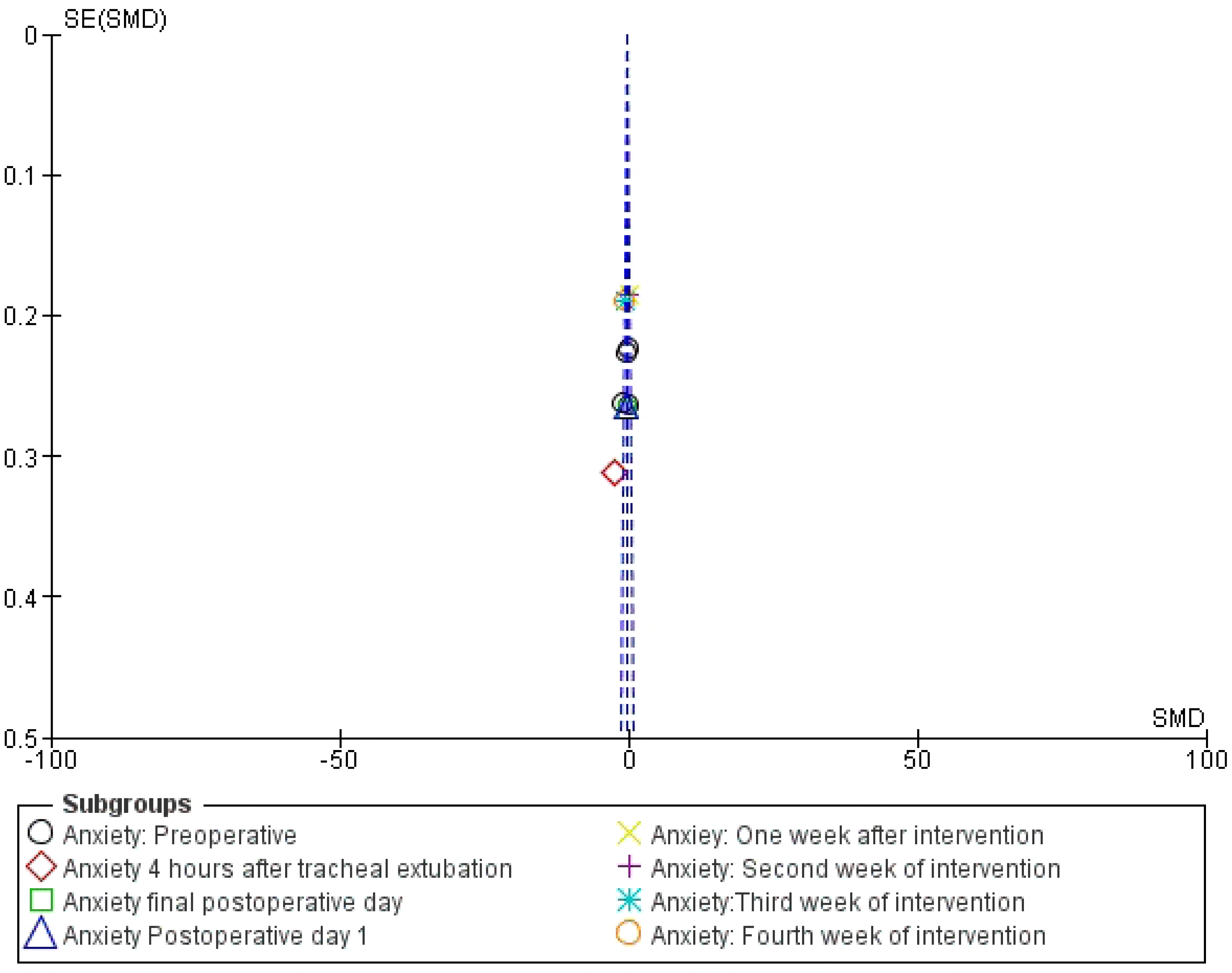
c. A funnel plot to assess publication bias for anxiety outcomes

The pooled overall effect of the combined therapy of aromatherapy and music on preoperative anxiety reduction was not significant whereas the effect on postoperative anxiety was statistically significant. On average, combined aromatherapy and music therapy reduced anxiety scores by 1.08 units (SMD = -1.08, 95 % CI: -1.45, -0.70), compared to the control condition. This estimate was driven from the study conducted by Deng et al., 2022, which assessed the effect of aromatherapy both preoperatively and postoperatively **Fig 6).** Comparing aromatherapy with music therapy, the difference was not statistically significant preoperatively, but postoperatively. Across time periods, the group exposed to aromatherapy had 0.39 (SMD=0.39, 95% CI 0.12,0.65) higher anxiety scores compared to music therapy indicating that music therapy is superior to aromatherapy in reducing anxiety (**Figure 7).**

**Figure 6.**
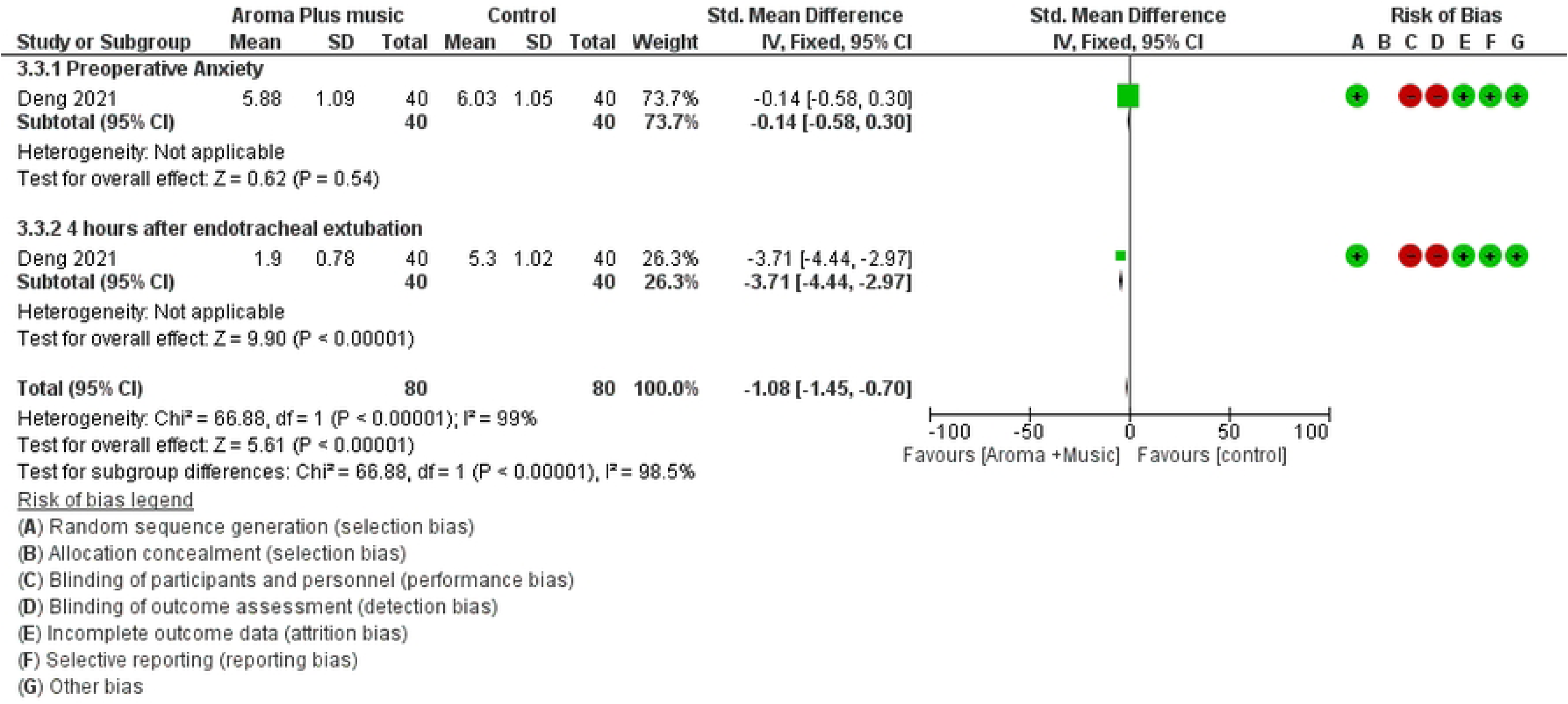
A forest plot comparing the effect of aromatherapy on pain scores to music therapy. A forest plot showing the effect of aromatherapy combined with music on anxiety scores compared to usual care.

**Figure 7.**
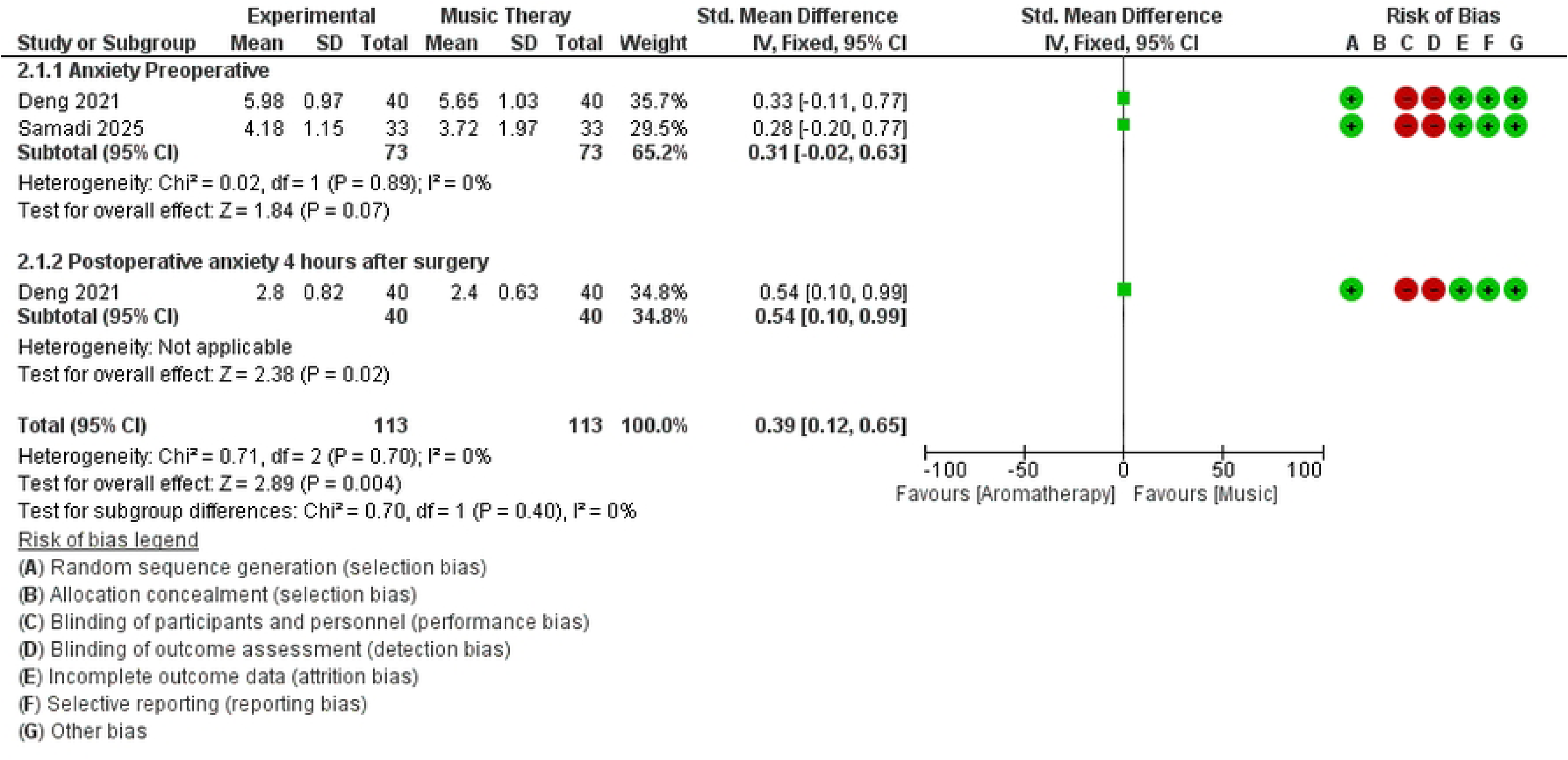
A forest plot comparing the effect of aromatherapy on anxiety scores to music therapy.

### Summary of findings

Certainty assessment was made using the GRADE methods for both outcomes.

#### Pain

On average, aromatherapy reduces pains scores by 0.79 units (SMD=-0.79, 95% CI - 1.42, -0.16) compared to control condition (Low-quality evidence) **(Table 5).**

**Table 5.**
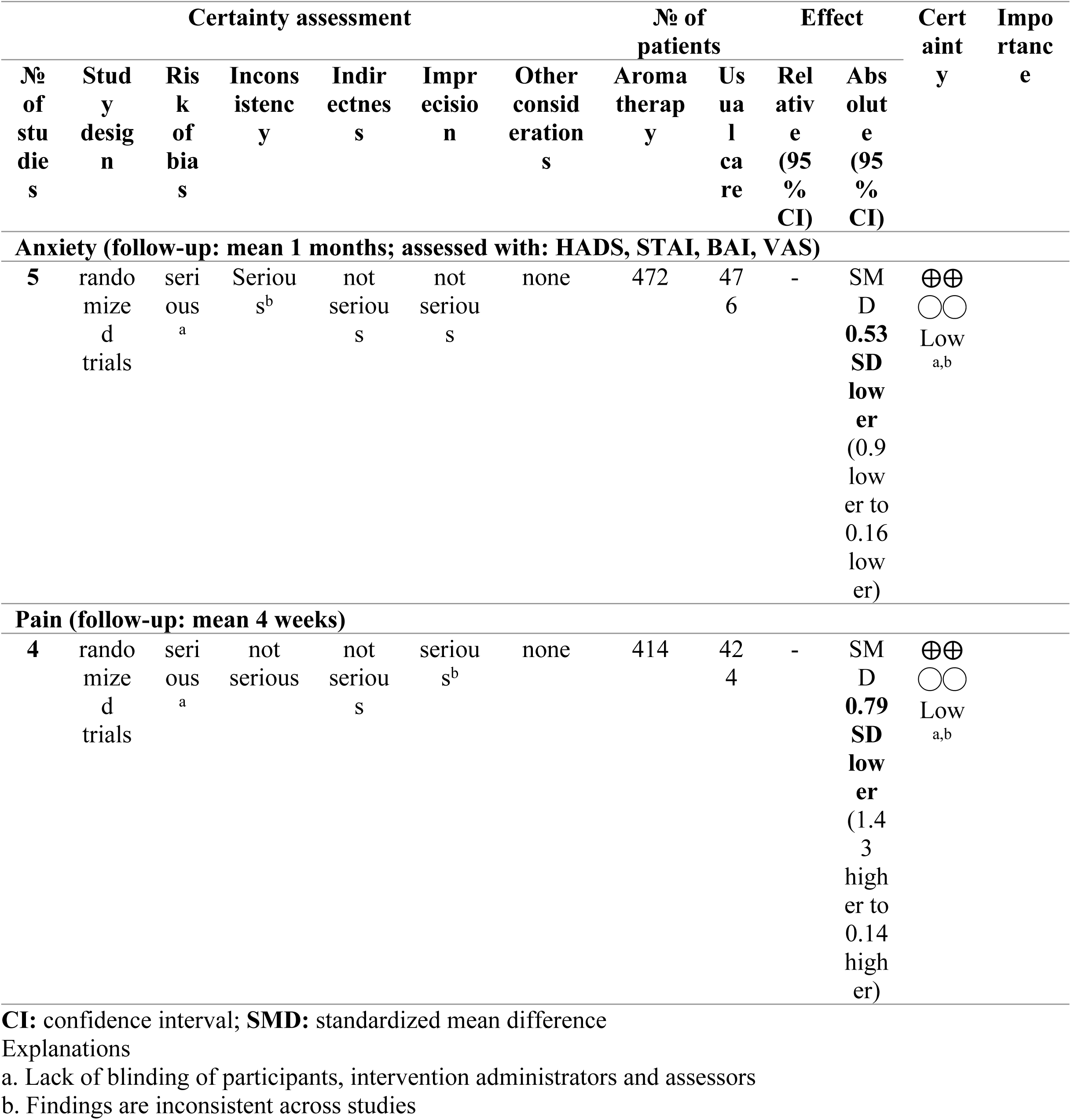
Summary of findings for the effectiveness of aromatherapy compared to Usual care for breast cancer patients for anxiety and pain management.

Overall, across the different time points, patients exposed to the combination of aromatherapy and music therapy had 1.26 (SMD= -1.26, 95 CI=-1.65, -0.87) units lower pain scores compared to usual care (Low-quality evidence) **(Table 6).**

**Table 6.** Summary of findings for the effectiveness of combined aromatherapy and music therapy for anxiety and pain management in breast cancer patients.

| Certainty assessment |  |  |  |  |  |  | № of patients |  | Effect |  | Certainty | Importance |
| --- | --- | --- | --- | --- | --- | --- | --- | --- | --- | --- | --- | --- |
| № of studies | Study design | Risk of bias | Inconsistency | Indirectness | Imprecision | Other considerations | Aromatherapy combined with music therapy | usual care | Relative (95% CI) | Absolute (95% CI) |  |  |
| Anxiety (follow-up: 4 hours) |  |  |  |  |  |  |  |  |  |  |  |  |
| 1 | randomized trials | serious<sup>a</sup> | serious<sup>b</sup> | not serious | not serious | none | 80 | 80 | - | SMD 1.08<br>SD lower (1.45 higher to 0.7 higher) | ⊕⊕<br>○○<br>Low<sub>a,b</sub> |  |
| Pain (follow-up: mean 4 weeks) |  |  |  |  |  |  |  |  |  |  |  |  |
| 1 | randomized trials | serious<sup>a</sup> | serious<sup>b</sup> | not serious | not serious | none | 80 | 80 | - | 1.26 SMD<br>lower (1.65 lower to 0.87 lower) | ⊕⊕<br>○○<br>Low<sub>a,b</sub> |  |

CI: confidence interval; SMD: standardized mean difference
Explanations
a. No blinding of participants, intervention administrators and outcome assessors
b. The effectiveness of intervention is not consistent across time points and studies.

There is no significant difference between aromatherapy and music therapy on both preoperative and postoperative pain scores (Very low-quality evidence) **(Table 7).**

**Table 7.**
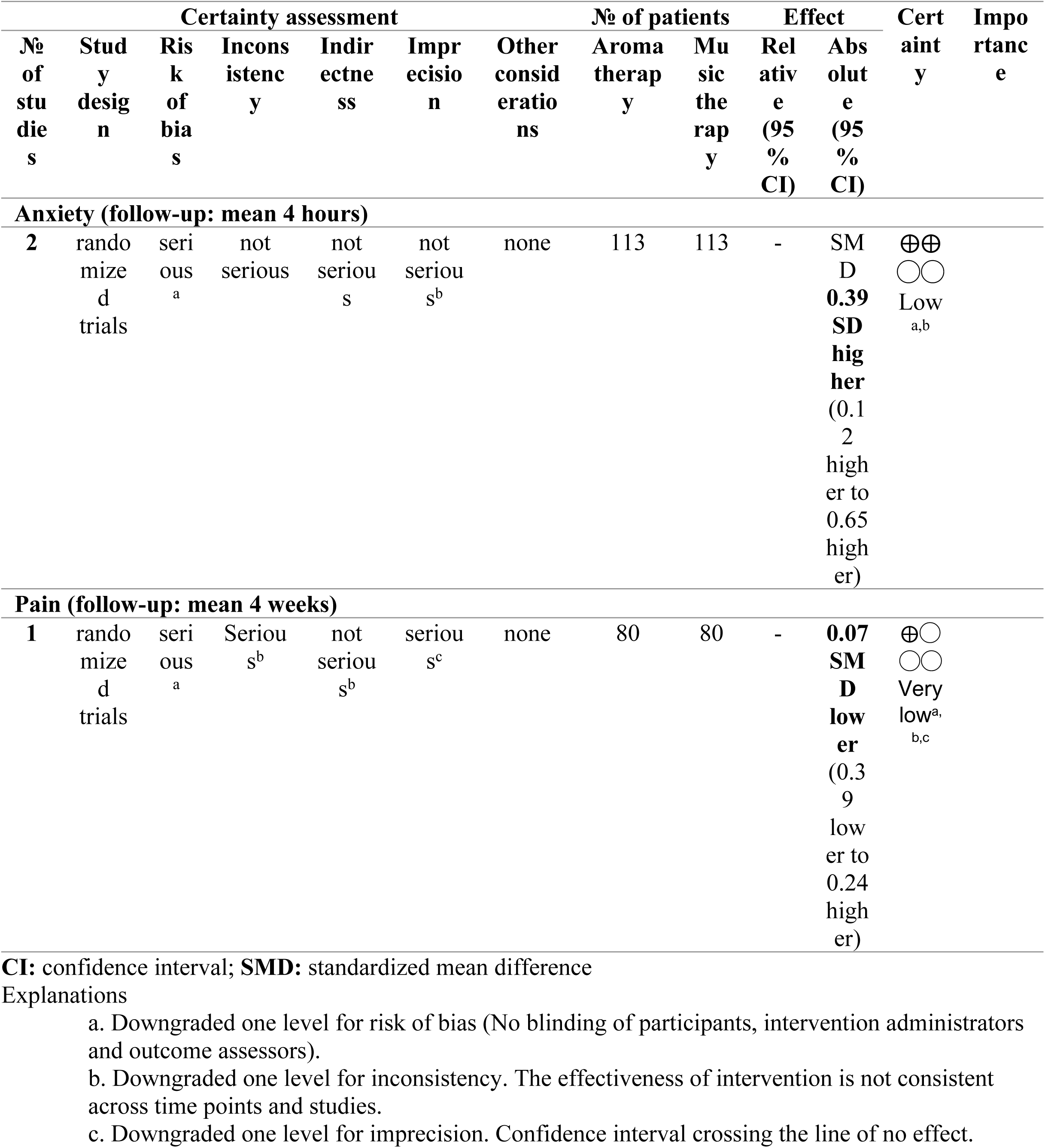
Summary of findings for the comparison of aromatherapy with Music therapy for anxiety and pain management in breast cancer patients.

#### Anxiety

Low-quality evidence indicates that on average aromatherapy reduces anxiety scores by 0.53 units (SMD=-0.53, 95 CI=-0.90, -0.16) units compared to standard care **(Table 5).**

Low-quality evidence indicates that on average the combination of aromatherapy and music therapy reduced anxiety scores by 1.08 units (SMD = -1.08, 95 % CI: -1.45, -0.70) units compared to standard care **(Table 6).**

Low-quality evidence indicates that aromatherapy’s effectiveness is inferior compared to music therapy in reducing anxiety scores. Aromatherapy group had 0.39 [SMD=0.39, 95% CI 0.12,0.65) higher units in anxiety scores compared to music therapy **(Table 7).**

## Discussion

### Overall findings

Overall, studies included in this systematic review reported that aromatherapy is effective in reducing anxiety scores, with inconsistent effect on preoperative anxiety. Across time points, on average, aromatherapy reduced anxiety scores by 0.53 units and pain scores by 0.79 units [Low-quality-evidence].

Unlike the study by Behzadmehr et al., 2020 (16), these results align with more recent systematic reviews, Ahn & Kim, 2024 (29); Li et al., 2022 (30), which demonstrated reductions in anxiety, especially following inhalation aromatherapy. This suggests the potential impact of aromatherapy to significantly reduce anxiety amongst breast cancer patients if used as part of routine care. However, there is no adequate evidence supporting the use of aromatherapy for preoperative pain and anxiety.

### Effects of aromatherapy on pain

On average, aromatherapy reduced pain scores by 0.79 units (Low-quality evidence). While the overall effect estimate is significant, as previously stated, reporting on pain reductions from the included studies was mixed with half of the studies demonstrating significant benefits. For instance, study by Ha et al., 2022 (27) reported no significant difference pain scores between aromatherapy lymphatic massage (ALT) group and control group (1.73 vs 1.92), (p = 0.216). The lack of statistically significant differences might be because the control group were managed using standard analgesics (taxane administration). Similar mixed results have been reported in a previous systematic review by Shin et al., 2016 (31), reporting that aromatherapy massage may provide more relief for pain though the quality of the studies were poor and unreliable. On the other hand, Ha et al., 2022 (27) found that breast cancer patients who previously experienced TAPS (taxane acute pain syndrome) reported lower VAS pain scores after receiving ALT indicating importance of aromatherapy even in the studies that failed to demonstrate statistically significant effects. The other study that reported statistically non-significant effect was Shammas et al., 2021 (26), which compared patients receiving lavender oil and those receiving placebo, coconut oil (p = 0.30). Again, the lack of statistically significant effect might be related to a comparable control group that was exposed to coconut oil. In addition, the study reported significant improvement in pain scores throughout the length of the patients’ hospitalization (26).

Interestingly, greater reduction in pain scores were recorded among studies that combined aromatherapy with the supportive interventions, music and massage, as opposed to aromatherapy alone (26). In addition, significant pain reduction was reported by aromatherapy alone in the study by Zhang et al., 2025 (17) which compared aromatherapy with sweet almond oil. Another important consideration is that aromatherapy was not effective on preoperative pain. This may be due to the short duration of time (30 minutes) between the initiation of the intervention and the assessment.

In general, aromatherapy has been found to alleviate pain in many studies of various patients which had been attributed to the application of essential oils to calm patients (32). On the other hand, there are few points that might be considered for cancer patients. Cancer pain is a symptom defined as the “simultaneous sensation of acute and chronic pain” that can be associated with multiple factors such as “invasive spread of tumor cells”, “consequence of treatments” and other intense sensations (33). Due to its multi-factorial nature, breast cancer pain management may need to include both physiological and psychological components, which makes it more difficult to address it with aromatherapy alone. Thus, its combination with massage and music therapy were shown to possibly be more effective in reducing pain. Nonetheless, lavender oil was found to be effective in two out of the three studies that examined its effectiveness in reducing pain. Linalool and linalyl acetate are chemical constituents of lavender that are believed to contribute to its therapeutic effect through inhibition of GABA(A) receptors in the central nervous system to induce a state of relaxation and mitigate pain perception (27, 34, 35). While aromatherapy is still useful compared to no interventions, there is no adequate evidence showing the superiority of aromatherapy to other pain management methods. Further studies may be needed to generate evidence on the effectiveness of aromatherapy across the different types of oils used.

### Effects of aromatherapy on anxiety

On average, aromatherapy reduced anxiety scores by 0.53 units [Low-quality-evidence]. Five of the six included studies reported statistically significant improvements in anxiety outcomes, particularly when lavender oil was administered. Most of the studies reported significant reductions regardless of whether aromatherapy was administered alone or in combination with other CAM therapies, except for one study which focused on aromatherapy alone by Shammas et al., 2021 (26). The reason behind the effectiveness of aromatherapy might be linked to the fact that there is direct connection between the olfactory stimulation and the limbic system, particularly the amygdala and hippocampus (36). Olfactory receptors can furthermore be expressed in non-olfactory tissues, such as the skin and the gastro-intestinal system (37), thus further implying the significance of aromatherapy message in the included study by Zhang et al., 2025 (17).

Most of the studies that reported on anxiety found significant reductions from the application of aromatherapy alone (75%), which aligns with the results from a recent meta-analysis that found that inhalation aromatherapy (alone) may help relieve symptoms of anxiety in cancer patients (37). The pooled findings from another previous meta-analysis of randomized controlled trials on the effects of aromatherapy on anxiety by Gong et al., 2020 (38) demonstrated that different kinds of oils can alleviate anxiety, with lavender essential oil showing the most beneficial results. This is consistent with the findings of the current review. These findings suggest that aromatherapy may provide meaningful psychological benefits when used alongside standard care for anxiety.

### Complementary and alternative medicine (CAM) utilization and integration of aromatherapy

An important consideration emerging from the findings in this systematic review is the role of multimodal interventions. Studies that combined aromatherapy with other non-pharmacological approaches, such as music therapy or massage, often reported greater improvements in both pain and anxiety outcomes compared to aromatherapy alone. For instance, the combination therapy group in the study by Deng et al., 2022 (25) demonstrated the largest reductions in anxiety and pain scores compared to the control group and therapies (aromatherapy and music) alone. This suggests a potential synergistic effect which highlights the importance of considering aromatherapy not as a standalone intervention, but as part of a broader integrative care strategy. Massage, in particular, may enhance the therapeutic effects of essential oils by increasing parasympathetic activity, improving circulation, and reducing nervous irritability through increased levels of dopamine and serotonin, which leads to relaxation and mood improvement (37).

### Strengths and limitations

Despite providing a comprehensive synthesis of existing evidence on the effectiveness of aromatherapy on pain and anxiety for breast cancer patients, some limitations must be considered while interpreting the findings reported in this review. Relatively fewer studies imply that there is a need for further studies. As this review was limited to English language articles, we might have missed non-English language articles. This is especially important as aromatherapy has been embedded in some cultures. Substantial heterogeneity observed among studies (I² = 89.3%) indicated considerable variability in study outcomes. However, through subgroup analysis was conducted based on timing of assessment, which revealed the timing of effectiveness (preoperative vs postoperative anxiety and pain). On the other hand, variations in essential oil used (e.g., lavender, rose, frankincense) highlighted the clinical variability of aromatherapy. This is significant as different neurotransmitters can be released depending on the type of aroma administered (36). Further research may clarify this.

On the other hand, this review is unique from other previous reviews in that it clearly indicated the distinction between the effects of aromatherapy itself and those of accompanying interventions, such as massage and music. When assessing the intervention alone, lavender essential oil was found to be the most common and beneficial oil for significant reductions in pain and anxiety. However, only one study (Deng et al., 2022 (25)) has demonstrated the possibility that significant reductions in anxiety in the combined therapy of music and aromatherapy can be attributed more to the effectiveness of music. Nevertheless, there is inconsistency across time (no significant effect on preoperative anxiety and pain but significant effect was demonstrated on post operative anxiety and pain. While there is inconsistency across time (no difference preoperatively), higher reductions of anxiety were obtained following music therapy post operatively compared to aromatherapy. It appears that music therapy is superior compared to aromatherapy (Low-quality evidence). There was no difference in pain scores between music therapy and aromatherapy (Very low-quality evidence). More research is required to confirm these findings.

The methodological limitations of the included studies further impact the strength of the evidence. Although most studies employed randomized controlled designs, issues such as the lack of blinding increase the risk of bias. The nature of aromatherapy interventions challenges the process of blinding, which may have led to expectancy effects among participants and researchers. As a result, the findings reported in some studies should be interpreted with caution.

There is a need for further studies to address the existing gap; specifically well-designed randomized controlled trials that will generate evidence on the effectiveness of aromatherapy on pain. Moreover, additional trials are needed to ensure greater consistency in the selection of essential oils, dosing protocols, and outcome measurement scales used. Furthermore, findings from this review underscore the need for future trials that directly compare aromatherapy alone versus combination therapies, for example aromatherapy plus massage or music therapy, to help clarify the extent to which aromatherapy independently contributes to observed therapeutic effects. Reviews of aromatherapy with massage indicate modest, transient benefits, but it remains unclear whether essential oils provide additional effects beyond massage alone. More research is also suggested to evaluate the effectiveness of music therapy in comparison to aromatherapy.

## Conclusion

### Implication for practice

The findings of this review suggest that aromatherapy may be effective in reducing anxiety and pain among breast cancer patients [Low-quality evidence]. Except for preoperative anxiety, the effect of aromatherapy on anxiety is consistent. While aromatherapy is consistently ineffective on preoperative pain, its effect on postoperative anxiety is inconsistent. The potential benefits of aromatherapy appear to be enhanced when used in combination with other complementary therapies such as massage and music. From a clinical perspective, aromatherapy offers low-cost, non-invasive CAM intervention with reported significant effects, making it an appealing complementary therapy for breast cancer patients. Given the promising findings on the effect of aromatherapy on pain and anxiety reported in this review, aromatherapy might be further considered as a complementary therapy after additional contextual considerations, especially for non-preoperative anxiety and pain.

### Implication for future research

Due to the variability of the specific aromatherapy used and the overall methodological and statistical heterogeneity, and the potential risks of bias related to the methodological limitations of the studies included in this review, further high-quality research is required to establish the efficacy of aromatherapy to better inform clinical practice. Future clinical trials might generate additional evidence of its integration with music therapy based on its apparent effectiveness in reducing anxiety and pain. This is particularly relevant in the context of breast cancer care, where psychological distress is commonly associated with pain-exacerbating non-adherence to treatment and worsening quality of life.

## Data Availability

All data for this manuscript have already been reported in the form of tables and figures.

## Acknowledgements

This project was partially supported by the American Cancer Society Cancer Research Institutional Grant DICRIDG-22-1012253-03-DICRIDG (Pilot Grant to GF) and the Gray Foundation. The work presented in the article is entirely that of the authors and was not influenced by the funders.

## Author contributions

Conceptualization: NOE, GTF and CLC.

Data curation: NOE, GTF.

Formal analysis: GTF.

Funding acquisition: GTF.

Investigation: NOE, GTF

Methodology: NOE, GTF and CLC.

Project administration: NOE, GTF and CLC.

Resources: GTF.

Software: NOE, GTF.

Supervision: GTF and CLC.

Validation: NOE, GTF and CLC.

Visualization: GTF and NOE.

Writing original draft: NOE.

Writing, review and editing: NOE, GTF and CLC.

## Availability of data, code and other materials

All data for this study has been reported in the form of tables and figures.

## Supplementary files

Supplementary file 1. PRISMA checklist.

Supplementary file 2. Search strategy.

